# Persistently increased systemic ACE2 activity and Furin levels are associated with increased inflammatory response in smokers with SARS-CoV-2 COVID-19

**DOI:** 10.1101/2021.01.14.21249836

**Authors:** Gagandeep Kaur, Shaiesh Yogeswaran, Thivanka Muthumalage, Irfan Rahman

## Abstract

**Background:** Tobacco smoking is known to be involved in the pathogenesis of several cardiopulmonary diseases, and smokers are susceptible to infectious agents. However, the progression of lung injury based on COVID-19 susceptibility and severity amongst smokers and those with pre-existing pulmonary diseases is not known. We determined the systemic expression and activity of COVID-19 related proteins, cytokine/chemokines, and lipid mediators (lipidomics) amongst COVID-19 patients with and without a history of smoking with a view to define biomarkers.

**Methods:** We obtained serum from COVID-19 positive and COVID-19 recovered patients with and without a history of smoking. We conducted a Luminex multiplex assay (cytokine levels), LC/MS (eicosanoids or oxylipin panel) and enzymatic activity assays on the serum samples to study the systemic changes in COVID-19 patients.

**Results:** On comparing the cytokine profiles among COVID-19 positive and COVID-19 negative patients, we found a significant upregulation in the production of pro-inflammatory cytokines like IL-1α, IL-8, IL-2, VEGF and IL-10 in COVID-19 positive patients as compared to the respective controls. Interestingly, smoking history resulted in further augmentation of the release of some hyper-inflammatory cytokines, like IFN-γ, Eotaxin, MCP-1 and IL-9 amongst COVID-19 positive patients. The enzymatic activity for ACE2, the binding partner for SARS-CoV2 virus in the host cell, was found to be significantly increased in the serum of patients with a smoking history compared to the serum collected from the non-smoking controls. Similarly to our cytokine analysis, our measurement of serum Furin levels was also affected by the patient’s smoking history, in which we reported a substantial rise in serum Furin levels of COVID-19 patients. The analysis of lipid mediators revealed a distinct signature amongst the COVID-19 positive versus recovered subjects in PGF_2α_, HETEs, LXA_4_ and LTB_4_ levels. However, we did not find any changes in the levels of any lipid mediators based on the smoking history of the patients. Overall, our results point towards distinct systemic signatures amongst COVID-19 positive patients. We also show that smoking adversely affects the systemic levels of inflammatory markers and COVID-19 related proteins, thus suggesting that COVID-19 infection may have severe outcomes amongst smokers which is reflected systemically.

## Introduction

The current pandemic of coronavirus disease 2019 (COVID-19) has emerged as a major public health threat worldwide. Viral pneumonia and acute respiratory failure are the most common clinical manifestations of severe COVID-19, featuring fever, cough, hypoxemia, dyspnea, and bilateral infiltrates on chest radiography (1). ACE2 (angiotensin converting enzyme 2) receptor is the viral binding site for the SARS-CoV2 in human. ACE2 is abundantly expressed in the lung epithelium, specifically type II pneumocytes, goblet cells, nasal epithelial/ciliated cells and oral mucosal cells (2-4). Under normal circumstances, ACE2 converts angiotensin 2 to metabolites, many of which exert vasodilatory properties. However, since the onset of this pandemic, ACE2 is being widely studied in the context of COVID-19. In this regard, it is still mostly unknown the specific nature of the change in ACE2 induced by SARS-CoV-2 infection, whether it’s the modulation of ACE-2 activity or its expression levels that have significance in affecting COVID-19 disease outcome.

With continual spreading of the virus and reports of a novel, mutant strain causing further alarm and panic, questions regarding COVID19 risk-factors have become even more urgent and a cause for concern. While old age, heart disease and diabetes are the universally accepted risk factors for COVID19, there are many other factors that are subject to debate and require empirical analysis. One such risk factor is smoking. While initial reports on COVID-19 risk factors have indicated little to no risk amongst smokers, recent data suggests otherwise (2, 5). A meta-analysis of 15 studies with a total of 2473 confirmed COVID-19 patients reported that COPD patients (63% vs 33.4 in people without COPD) and current smokers (22%) are at a higher risk of severity and mortality due to COVID-19 compared to disease-free and non-smoking individuals respectively (95% confidence interval)(6, 7). Similar findings were reported by Patanavanich et al (2020), whose research suggested that smoking nearly doubles the rate of COVID-19 progression amongst patients (8). Despite these findings, the exact pathogenesis of COVID-19 and the clinical features resulting in severe outcomes amongst smokers is largely unexplored.

In consideration of this lack of research and the growing call of concern for further research into COVID-19 risk-factors, we investigated whether a variation in systemic markers for inflammation and COVID-19 infection existed between smokers and non-smokers. We also studied the gender-based differences in the expression and activity of COVID-19 related proteins (ACE2 and Furin) to understand the progression of the disease amongst both sexes. Evidence from previous literature has suggested upregulated levels of ACE-2 in the lungs of smokers. However, there is no evidence correlating this increased expression to COVID-19 disease development and severity. Our study investigates the relationship between COVID-19 biomarkers and disease severity and shows a marked increase in the inflammatory spillover amongst COVID-19 positive patients with a smoking history compared to controls. We were also able to show a pronounced increase in cytokine/chemokine and Furin (S1/S2 cleavage protein) levels in serum of COVID-19 positive patients compared to the COVID-19 recovered controls. Lipid profiling further showed noticeable increases in the levels of Prostaglandin 2α, leukotrienes B_4_, lipoxin A_4_ and 15-Hydroxyeicosatetraenoic acid in serum collected from COVID-19 positive patients.

## Materials and Methods

### Ethics/Approval

All the procedures performed in this study comply with the protocols approved by the Institutional Review Board /Research Subject Review Board (RSRB) committee at the University of Rochester Medical Center, Rochester, NY with an approval number CR00002635. The patient samples and information used in this study were procured from a commercial provider-BioIVT (Westbury, NY, USA). All the laboratory procedures were performed in accordance with the regulations specified by the BSL2+ level of containment for Clinical and Research Safety.

Human study protocol: Yes; Animal study protocol: None; Institutional biosafety approvals: Yes. The University of Rochester Institutional Biosafety Committee approved the study (study approval number: Rahman/102054/09-167/07-186; identification code: 07-186)

### Human Blood serum collection

Sera from COVID-19 positive and COVID-19 recovered patients were obtained from BioIVT (Westbury, NY, USA). The patient population was categorized based on smoking status; both current and previous smokers were considered ‘Smokers’ for subsequent analyses. The characteristics of the study subjects used for the experiments are presented in **Table 1**. In this study, the results obtained from the sera of the COVID-19 recovered patient group are considered equivalent to COVID-19 negative patients.

**Table 1:** Characteristics of COVID-19 positive and negative subjects.

| Characteristics | COVID-19 negative patients | COVID-19 positive patients |
| --- | --- | --- |
| N | 21 | 16 |
| Age (years), mean (SD) | 46.71 (13.44) | 39.75 (14.68) |
| Smoker, n (%) | 7 (33.33) | 3 (18.75) |

### Assessment of pro-inflammatory mediators in blood sera using Luminex multiplex assay

The levels of pro-inflammatory cytokines/chemokines like MCP-1, IL-8, IFN-γ, TNF-α and IL-7 in the sera were measured by Luminex multiplex assay using Bio-Plex Pro™ Human cytokine 27-plex assay (Cat#M500KCAFOY, BIO-RAD, Hercules, CA) as per manufacturer’s directions. Blood plasma was diluted two-folds and the levels of 27 pro-inflammatory mediators (expressed as pg/ml) were measured using Luminex FlexMap3D system (Luminex, Austin, TX).

### Assessment of Furin levels using ELISA

To determine the level of Furin in sera collected from COVID-19 positive and negative patients, we employed Human Furin ELISA kit (Cat #: ab113322, Abcam, Cambridge, MA) as per the manufacturer’s protocol. Colorimetric detection was performed at 450 nm using Cytation 5 microplate reader (BioTek Instruments, Inc. Winooski, VT). Furin levels were expressed as pg/ml.

### Assessment of ACE2 Activity

We utilized the ACE2 Activity Assay kit (Cat #: K897 BioVision, Milpitas, CA, USA) to determine the ACE2 activity in the human serum samples. The assay was performed as per the manufacturer’s instructions. In brief, serum samples were lysed through the addition of equal volumes of ACE2 Lysis Buffer and ACE2 Assay Buffer. The lysed serum samples, in addition to the appropriate standards and controls (positive, negative, and background), were then added to a 96-well plate. After that, 50 ul of ACE2 substrate was added to both sample and control wells, and subsequently, fluorescence was measured at an excitation maximum of 320 nm and an emission maximum of 420 nm using Cytation 5 microplate reader (BioTek Instruments, Inc. Winooski, VT). Total protein content per sample was determined using the Bradford protein assay kit (Thermo Fisher, Waltham, MA). Sample ACE2 activity for each sample was calculated using the following formula:

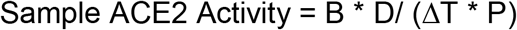

where, B = Released MCA (cleaved product of ACE2 substrate) in Sample based on standard curve slope, ΔT = Reaction time (in min), P = Sample used (in mg), and D = Sample Dilution factor.

### Determination of Serum Eicosanoid/Oxylipins levels through Lipidomic Analysis

Serum eicosanoid/oxylipin profiling was outsourced to and performed by Cayman Chemical (Ann Arbor, MI). Lipid profiling was done using ultraperformance liquid chromatography in tandem with mass spectroscopy (UPLC-MS/MS). Lipidomes were prepared using serum from six different patients from each group.

### Nomenclature

The abbreviations used for various classes of lipids include the following: 6-keto PGF1α: 6-keto Prostaglandin F_1a,_ TXB2: Thomboxane B_2,_ PGF_2α_: Prostaglandin F_2α_, PGE2: Prostaglandin E_2_, 12-HHTrE: 12-Hyrdoxyheptadecatrenoic acid, LTB4: Leukotriene B_4_, LXA4: Lipoxin A_4_, HETE: Hydroxyeicosatrtraenoic acid, EET: epoxyeicosatrienoic acid, HODE:Hydroxyoctadecadienoic acid and HDHA: Hydroxy Docosahexaenoic Acid.

### Lipid extraction

Lipid extraction from the serum samples was performed by protein precipitation followed by solid-phase extraction (SPE). Protein precipitation was performed by addition of 50 µL H2O: Acetonitrile solution to each sample. Thereafter, SPE was performed using Strata-X cartridges (33 μm, 200 mg/10 mL; Phenomenex, PA). The extracted lipids were finally dissolved in 100 µL water/acetonitrile 60:40 (v:v) solution. To prepare the calibration curves, a mixture of the 20 calibration standards was prepared in methanol.

### UPLC-MS/MS

10 ul of calibration standards and samples was added to Kinetex (2.6 µm C18 100 Å 100×2.1 mm, Phenomenex OOD-4462-AN) column and Reverse phase liquid chromatography (LC) using Sciex ExionLC Integrated System was used for lipid separation. The lipid quantification in the samples was performed using Sciex 6500+. The total amount of eicosanoids present in each sample was determined using MultiQuant software (Sciex). The lipid abundance ratios were calculated in terms of log base 2-fold change and plotted as a heat map.

### Statistical Analyses

All statistical calculations were performed using GraphPad Prism 8.0. Data is expressed as mean ± SE. Pairwise comparisons were done using unpaired Student’s *t*-test. Differences were considered statistically significant at ∗p < 0.05, ∗∗p *<* 0.01, and ∗∗∗ p *<*0.001 when compared with respective controls.

## Results

### ACE-2 activity varies as function of smoking history among COVID-19 patients

Since in humans, ACE2 binds to the SARS-CoV2 spike protein, we investigated changes in COVID-19 positive and COVID-19 negative serum ACE2 activities. To our surprise, we found a significantly higher ACE2 activity in serum from COVID-19 negative patients compared to serum from COVID19 positive patients. **(Fig 1a)**. This could have occurred as in this study the COVID-19 negative patient sera actually belong to COVID-19 recovered patient group. As expected, after repeating these experiments on blood plasma samples from COVID-19 positive and healthy subjects, we found a significant increase in ACE2 activity in the COVID-19 positive samples compared to healthy controls (data not shown).

**Figure 1:**
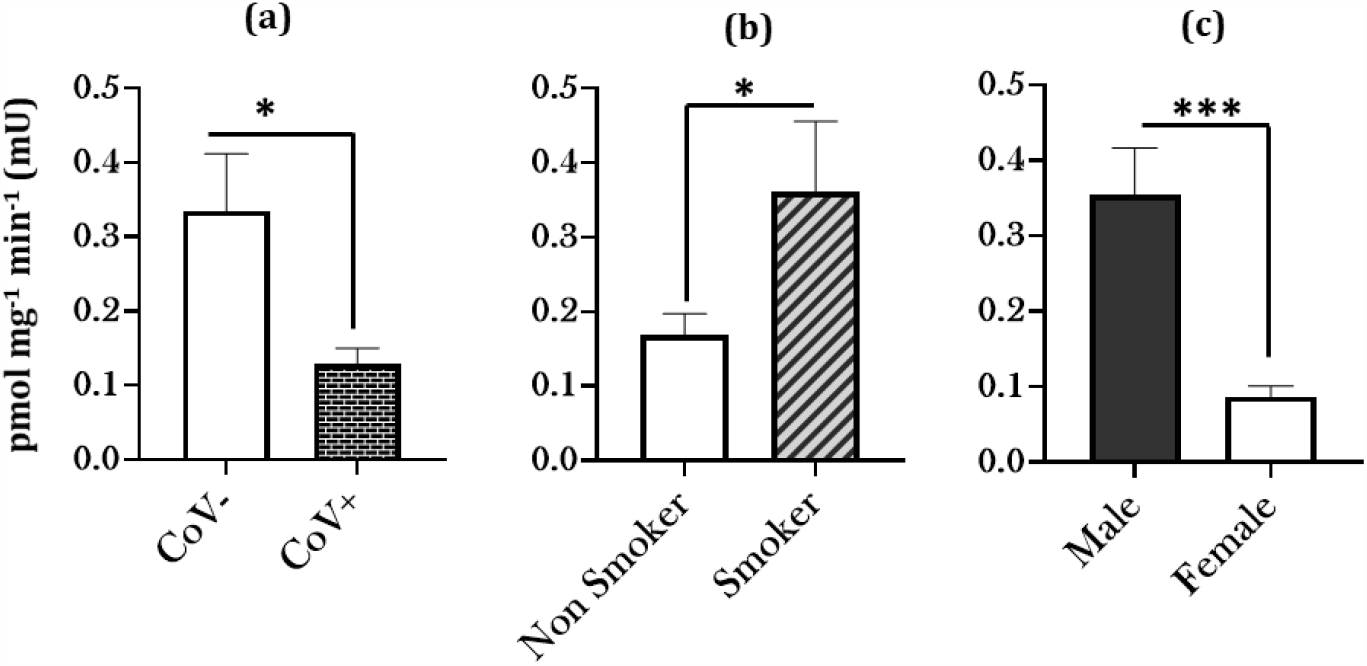
Increased ACE2 activity amongst COVID-19 patients with a smoking history. Patient Sera from COVID-19 positive and COVID-19 negative (COVID19 recovered) subjects was obtained and the ACE2 activity was quantitatively measured. The obtained results were plotted based on serum Furin levels in (a) COVID-19 negative (COVID-19 recovered) vs COVID-19 positive patients, (b) COVID-19 positive (current or previous) patients with or without a smoking history, and (c) COVID-19 patients based on their gender. Data are shown as mean ± SEM (n = 8–15/group). ∗p<0.05, ∗∗∗ p<0.0001 as per Student’s t-test for pairwise comparisons. CoV-: COVID-19 negative (COVID-19 Recovered), CoV+: COVID-19 positive patients.

However, it was interesting to find a significant increase in ACE2 activity in serological samples from COVID-19 patients (current and recovered) with a smoking history as compared to non-smokers **(Fig 1b)**. In fact, ACE2 activity was found to be more pronounced among male patients compared to females **(Fig 1c)**. Our results show that age and smoking status play a crucial role in governing the COVID-19 related enzyme activity in human subjects, thereby affecting the disease pathogenesis amongst individuals.

### Smoking Upregulates Furin Expression in serological samples from COVID-19 patients

Another key to understanding COVID-19 virulence as a function of susceptibility to viral entry is analyzing changes in Furin levels. Unlike other Coronaviruses, SARS-CoV-2 has a lessened dependence on target host cell proteases and depends more on proprotein convertase Furin for its viral entry. Based on this, we measured Furin-levels in patient sera from COVID-19-positive and COVID-19 negative (COVID-19 recovered) groups using ELISA. We observed a marked increase in the serum Furin levels (p=0.083) in COVID-19 positive patients as compared to controls **(Fig 2a)**. A significant upregulation of Furin levels was noted among smokers compared to non-smokers among sera collected from COVID-positive (current and previous) patients **(Fig 2b)**. Similarly, although not significant, Furin levels among male patients were elevated compared to females; this suggests gender-based variations in the Furin expression on SARS-CoV2 infection. **(Fig 2c)**.

**Figure 2:**
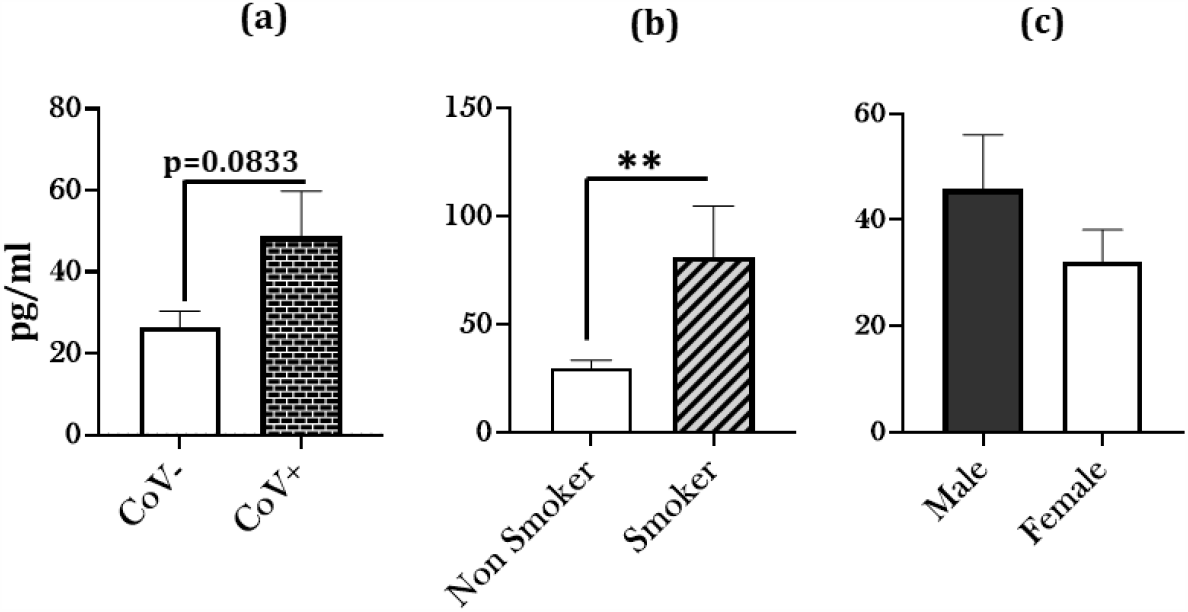
Increased Furin levels amongst COVID-19 patients with a smoking history. Blood serum from COVID-19 positive and COVID-19 negative (COVID-19 recovered) patients were obtained and the Furin levels were quantitatively measured. The obtained results were plotted based on serum Furin levels in (a) COVID-19 negative vs COVID-19 positive (COVID-19 recovered) patients, (b) COVID-19 positive patients (current or previous) patients with or without a smoking history, and (c) COVID-19 patients based on their gender. Data are shown as mean ± SEM (n = 8–15/group). ∗∗ p<0.001 as per Student’s t-test for pairwise comparisons. CoV-: COVID-19 negative (COVID-19 Recovered), CoV+: COVID-19 positive patients

### Infection with SARS-CoV-2 upregulates pro-inflammatory cytokine expression in smokers

It is well known that the exacerbation of the COVID-19 symptoms is associated with an increased expression of pro-inflammatory mediators. Given this, we analyzed the expression of 27 cytokines/chemokines in the serum samples from, COVID-19-positive and COVID-19-negative (recovered) patient population using Luminex multiplex assay. Our results showed significant increases in the levels of IL-8, IL-10, IL-2, VEGF and IL-1α (p= 0.0616) in COVID-19 positive patient sera compared to healthy controls **(Fig 3)**. Parameters such as, IL-5, GM-CSF, IL-12(p70), and IL-15 were undetected in the patient sera from diseased and normal subjects.

**Figure 3:**
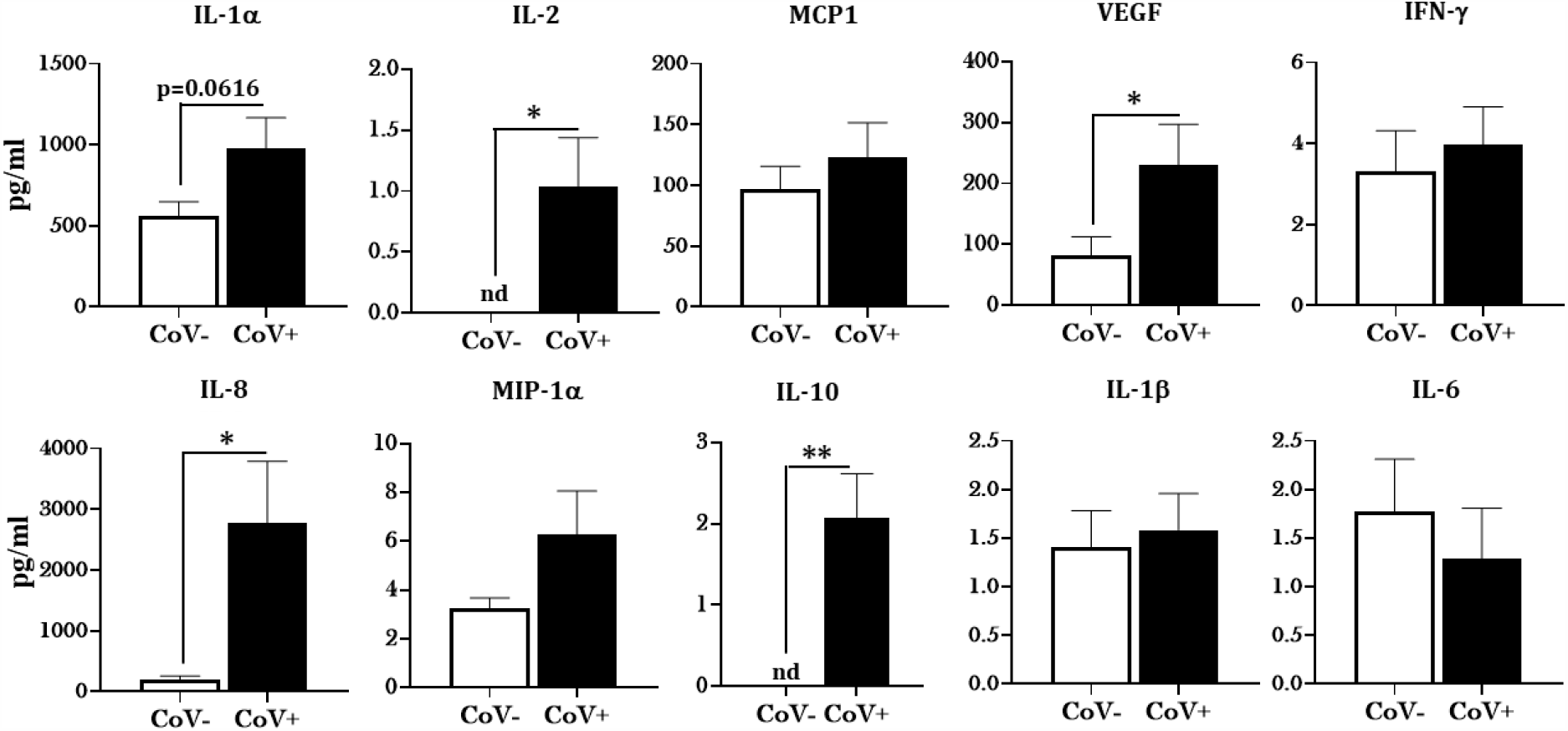
Elevated cytokine levels in COVID-19 positive patients. Blood serum samples from COVID-19 positive, and COVID-19 negative (COVID-19 recovered) patients were collected and the levels of cytokines/chemokines was assessed with the help of Luminex multiplex assay. The levels of detected cytokines was plotted using GraphPad Prism 8. Data are shown as mean ± SEM (n = 10-15/group). ∗ p<0.05, ∗∗ p<0.01; vs CoV-patients as per One-way ANOVA for multiple comparisons. CoV-: COVID-19 negative (COVID-19 recovered), CoV+: COVID-19 positive patients. Cytokines not detected IL-5, GM-CSF, IL-12(p70) and IL-15

Intriguingly, when analyzing cytokine/chemokine levels in patient sera based on smoking status, a unique trend emerged. We noted a substantial increase in the production of pro-inflammatory markers like, IFN-*γ* (p=0.0836), MCP-1 and Eotaxin in the COVID-19 positive patient sera who had a smoking history compared to the non-smoking controls.

Furthermore, we found a moderate increase in the levels of IL-9 (p=0.0991) amongst smokers infected with COVID 19compared to non-smokers **(Fig 4)**. These results support our hypothesis and further elucidate the role smoking has in exacerbating both disease severity and outcome with regards to SARS-CoV-2 virulence amongst smokers.

**Figure 4:**
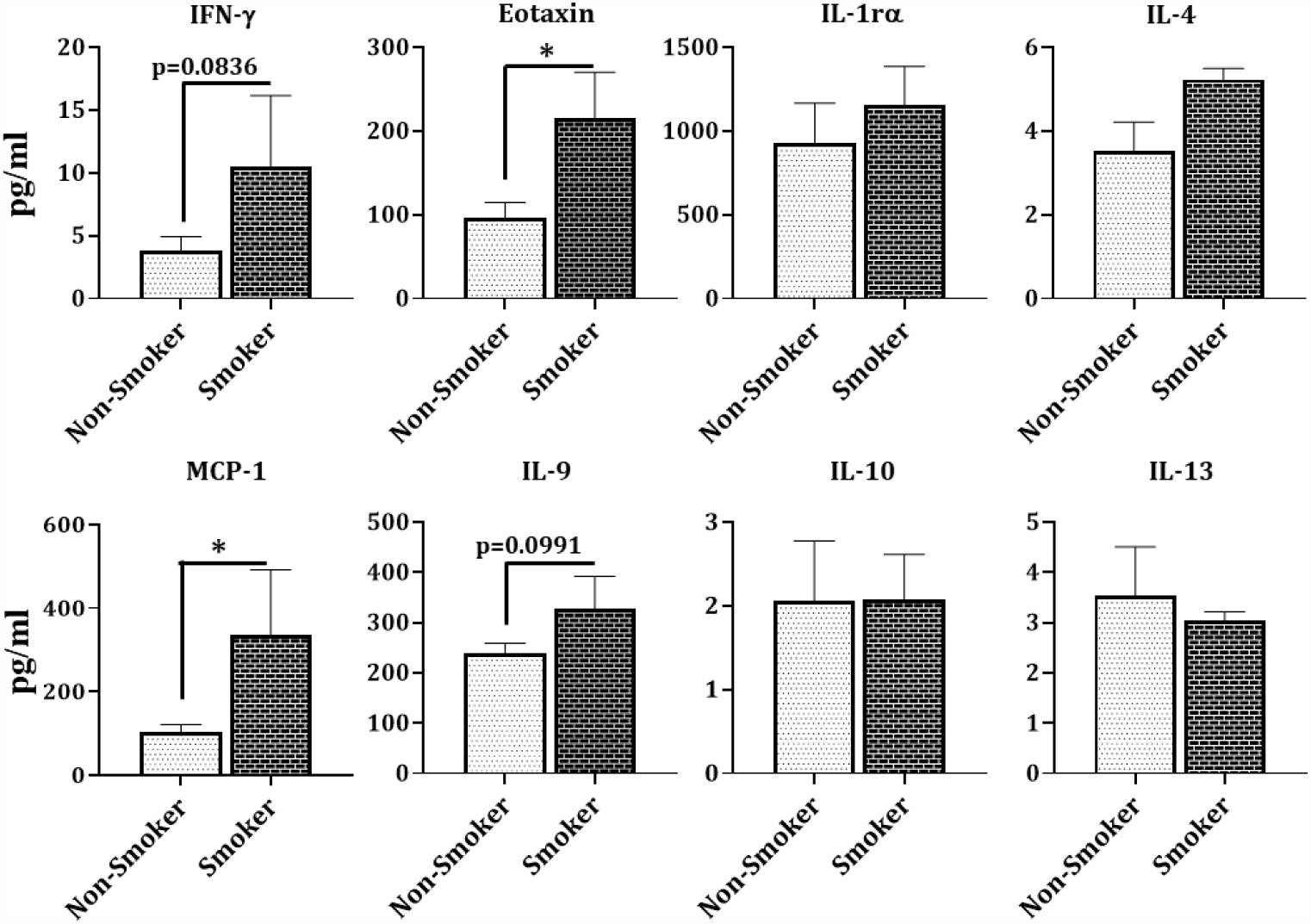
Hyperinflammation in COVID-19 positive patients with a smoking history. Levels of cytokines/chemokines as assessed with the help of Luminex multiplex assay in blood serum samples from COVID-19 positive patients with and without a smoking history (NS: non-smokers and smokers) were collected (n = 3–10/group). Data are shown as mean ± SEM. ∗p<0.05, as per Student’s t-test for pairwise comparisons.

### Altered serum lipid profile amongst COVID-19 positive patients

It is known that infections can induce a variety of alterations in lipid metabolism that 4could dampen inflammation or fight infection. Thus, we were next interested in studying the changes in the lipid profiles of patient sera from COVID-19 positive and COVID-19 negative groups. **Fig 5** depicts a generated heat map which shows alterations in the levels of the 18 most prevalent eicosanoids/oxylipins in sera from COVID-19 positive subjects compared to their respective controls. Most notably, we found noticeable increases in the levels of PGF_2α_, LTB4, LXA4, 5HETE, and 20-HETE in COVID-19 positive sera. The detailed account of the fold changes in the levels of each of the studied lipids is provided in **Table 2**.

**Table 2:** Differentially altered lipid analytes in COVID-19 positive patient sera with respective fold changes.

| Analyte | Log2 fold change<br>(Cov+ vs CoV-) | SD |
| --- | --- | --- |
| 6-keto PGF<sub>1a</sub> | 0.078348 | 0.483034 |
| TXB<sub>2</sub> | -0.35068 | 2.271691 |
| PGF<sub>2a</sub> | 0.854693 | 0.877836 |
| PGE<sub>2</sub> | 0.685295 | 1.65742 |
| 12-HHTrE | -0.56546 | 1.837558 |
| LTB<sub>4</sub> | 1.31646 | 1.100475 |
| LXA<sub>4</sub> | 0.112419 | 1.132973 |
| 5-HETE | 1.446217 | 1.704243 |
| 11-HETE | 0.948088 | 1.31254 |
| 12-HETE | 0.473277 | 0.935641 |
| 15-HETE | 0.808158 | 1.10055 |
| 20-HETE | 0.687371 | 0.892525 |
| 5(6)-EET | 1.123827 | 0.756739 |
| 8(9)-EET | -0.15191 | 3.07822 |
| 11(12)-EET | 0.714846 | 1.34743 |
| 9-HODE | -0.83099 | 1.994469 |
| 13-HODE | -0.61875 | 1.361414 |
| 17-HDHA | -0.7935 | 1.346715 |

**Figure 5:**
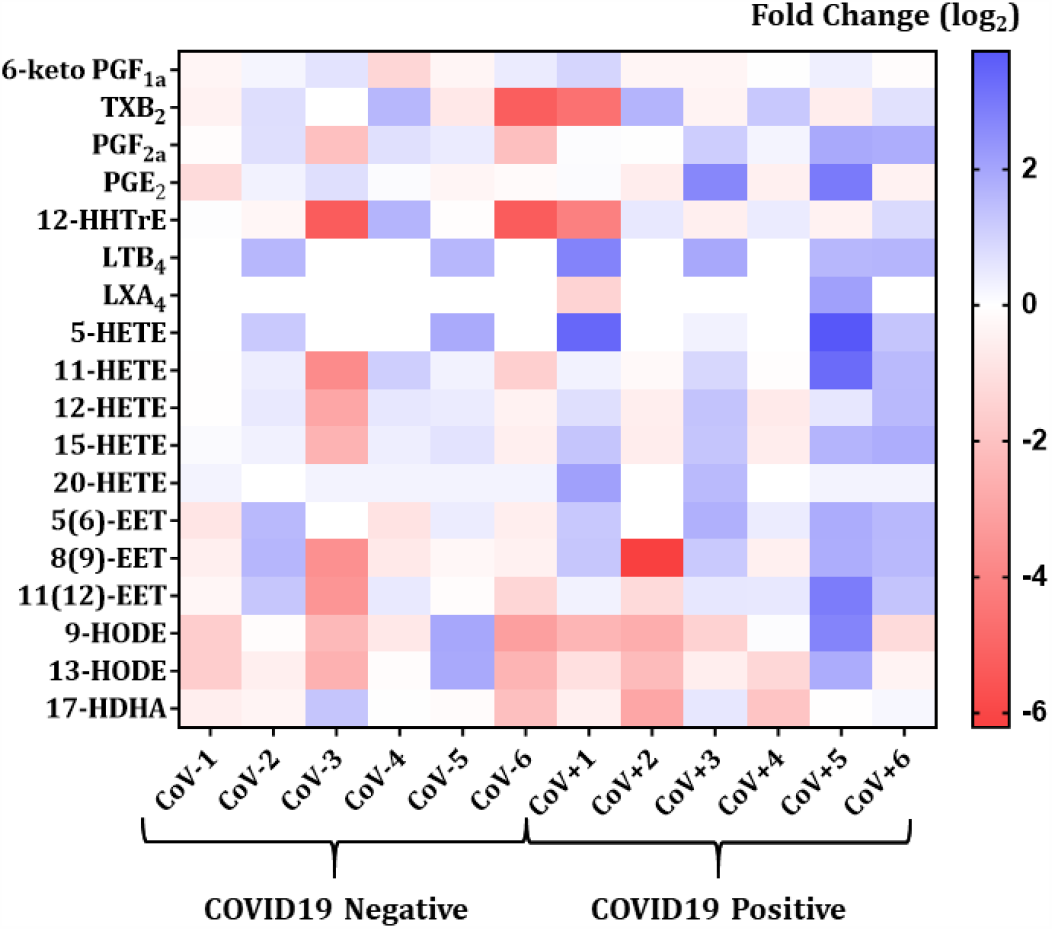
Heat map of eicosanoid levels in COVID-19 positive and COVID-19 negative patient serum. The fold changes in the eicosanoid levels in COVID-19 negative (COVID-19 recovered) patient serum as compared to COVID-19 positive serum. Each horizontal row represents the analyte in a specific lipid class and each vertical column represents the individual sample being tested. Lipid abundance ratios are colored according to the fold changes and the color key indicates the magnitude of log2 fold change.

## Discussion

The current pandemic of COVID-19 poses a serious threat to the global health and economy. While the efforts of rolling out an effective vaccine are ongoing, there is panic and uncertainty with regards to the new mutant strain of SARS-CoV2 (9). Amidst of this, it is important to recognize the high-risk populations and the underlying differences in the disease pathogenesis to limit the viral spread in an efficient manner. In this regard, we were interested in understanding the COVID-19 disease severity and progression amongst smokers and investigated the systemic responses against SARS-CoV2 infection in COVID-19 positive and COVID-19 negative patients.

We first investigated the ACE2 enzyme activity in the patient serum to determine the role of ACE2 in regulating SARS-CoV2 infection. ACE2 is well-known as the binding receptor for the SARS-CoV2 on host’s cell surface and use of ACE inhibitors and ARB blockers in COVID-19 patients is being debated since the start of the pandemic. To our surprise, we found an increased ACE2 activity in our COVID-19 negative patients as compared to COVID-19 positive individuals. It is pertinent to mention here that for this study we are denoting COVID-19 recovered patient sera as COVID-19 negative, which could be the reason for this observation. On determining the ACE2 activity in the plasma samples from COVID-19 positive and healthy individuals from a different cohort we observed a significant increase in ACE2 activity in plasma from COVID-19 positive patients as compared to the normal subjects **(Kaur et al**., **data not shown)**. This is an interesting find as it shows that ACE2 activity remains persistently elevated even after recovery and might have important implications for future research. Importantly, ACE2 is expressed in various tissues including the heart, kidney, lungs; and can undergo shedding into the blood circulation. Reports suggest that circulating ACE2 activity is indicative of adverse cardiovascular outcomes in patients with hypertension, coronary artery disease and aortic stenosis (10-12). It is also known to have a negative correlation with the glomerular filtration rate in type 1 diabetes (13, 14). Considering this, our results suggest dysregulated shedding of active ACE2 even after recovery amongst COVID-19 patients which may have adverse effects in the long run and is subject to further investigation.

Interestingly, we found that the ACE2 activity was significantly higher for individuals with a smoking history, thus suggesting that smoking can greatly affect the disease severity and outcomes amongst COVID-19 patients. As has been reported previously (15), our data also shows gender-based upregulation of ACE2 activity amongst males as compared to females. This could correlate to the increased morbidity and mortality amongst male patients and must be interesting to study in future.

We next studied the levels of Furin proteases in the sera of COVID-19 positive and COVID-19 negative patients. Furin enables the cleavage of S-protein thus facilitating viral cell entry into the host (16). Evidence suggests that furin cleavage plays a potent role in the virulence of dengue, HIV and avian flu (17, 18). We observed increased levels of serological Furin in COVID-19 positive patients as compared to controls. We further found that the levels of furin was significantly increased amongst smokers which substantiates our hypothesis that smoking is a risk factor for SARS-CoV2 infection.

Coinciding with the existing literature (19-21) we also found increased levels of cytokines/chemokines in the patient serum from COVID-19 positive subjects. Additionally, we for the first time, show significant changes in the levels of these pro-inflammatory mediators in COVID-19 positive patients with a smoking history as compared to the non-smoking controls. Upregulation in the factors like IFN-γ, MCP-1 and Eotaxin points towards increased severity of the disease amongst the smokers. Of note, we also found some changes in the lipid profiles of COVID-19 positive and COVID-19 negative patients. Viral infections are known to cause changes in lipid metabolism and play a crucial role in regulating innate and adaptive immune responses. Amongst the lipids that showed marked increase in COVID-19 positive patients were PGF_2α_, LTB_4_, LXA_4_, 15-HETE and 20-HETE. Of these, PGF_2α_, LTB_4_ and15-HETE are indicative of bronchoconstriction and lung injury (22-24).

Overall, our results show evidence of inflammatory spill-over in COVID-19 positive patients which is shown to be aggravated in patients who smoke. Pulmonary conditions like COPD and smoking-induced lung injury are known to cause such spillovers into the systemic circulation (25, 26). Thus, it will be important to develop these inflammatory and lipid mediators as biomarkers to ascertain disease severity amongst COVID-19 patients to provide rapid and effective care for better recovery. Though we have not discussed here but vaping population might be yet another population group that may suffer from severe outcomes in event of a SARS-CoV2 infection. Previous work by our group has shown gender-based variation in the ACE2 expression in lung tissues from C57Bl/6J mice exposed to e-cigarette (e-cig) aerosols (27). E-cig use has been associated with loss of lipid homeostasis eventually leading to pulmonary toxicity and lung injury (28, 29). Hence it is important to study the disease progress and pathogenesis in this patient population in smokers and vapers (e-cigarette users) in the future.

Though we were able to show variations in systemic inflammatory and lipid mediators on SARS-CoV2 infection, our study had some limitations. The sample cohort used for this study was relatively smaller and comprised of a relatively homogenous demographics. It is important to conduct these experiments on a larger sample population with a heterogeneous demographics to deduce better conclusions. Also, we intend to use patient serum from healthy or COVID-19 negative individuals in the future to compare the variations amongst COVID-19 positive, recovered and healthy subjects. The persistently elevated ACE2 activity in the COVID-19 recovered patients provides evidence for a much thorough investigation into the long-term health effects of SARS-CoV2 infection in smokers. Such an exploration is crucial to understand the mechanistic role of intact and circulating ACE2 in COVID-19 and deduce if recombinant ACE2 could develop as a therapy.

In conclusion, we show that the systemic ACE2 activity, Furin levels and cytokine release is upregulated amongst COVID-19 patients with a smoking history, thereby rendering them more susceptible to severe symptoms and disease outcomes. We also show that smoking adversely affects the systemic levels of inflammatory markers and COVID-19 related proteins, thus suggesting that COVID-19 infection may have severe outcomes amongst smokers which is reflected systemically. We also provide evidence for inflammatory systemic spillover due to COVID-19 which could be crucial in identifying biomarkers or developing future therapies, and/or monitoring therapies in susceptible population.

## Data Availability

All are Included

## List of Abbreviations

COVID-19: Coronavirus Disease-2019
ACE2: Angiotensin converting Enzyme 2
GM-CSF: Granulocyte-macrophage Colony stimulating Factor
HIV: Human Immunodeficiency Virus
COPD: Chronic obstructive pulmonary disease

## Acknowledgements

We thank Dr. Shikha Sharma for technical assistance.

## Declarations

The authors have declared that no competing interests exist.

## Author’s Contribution

GK, and SY designed and conducted the experiments, GS, SY, and IR wrote and edited/revised the manuscript. TM analyzed the lipidomic data and edited the manuscript. IR obtained research funding and conceptually designed the overall manuscript.

